# Rapid Systematic Review Exploring Historical and Present Day National and International Governance during Pandemics

**DOI:** 10.1101/2020.07.07.20148239

**Authors:** Elizabeth Lowry, Henock Taddese, Leigh R. Bowman

## Abstract

**Introduction:** Pandemics have plagued mankind since records began, and while non-communicable disease pandemics are more common in high-income nations, infectious disease pandemics continue to affect all countries worldwide. To mitigate impact, national pandemic preparedness and response policies remain crucial. And in response to emerging pathogens of pandemic potential, public health policies must be both dynamic and adaptive. Yet, this process of policy change and adaptation remains opaque. Accordingly, this rapid systematic review will synthesise and analyse evaluative policy literature to develop a roadmap of policy changes that have occurred after each pandemic event, throughout both the 20th and 21st Century, in order to better inform future policy development.

**Methods and Analysis:** A rapid systematic review will be conducted to assimilate and synthesise both peer-reviewed articles and grey literature that document the then current pandemic preparedness policy, and the subsequent changes to that policy, across high-, middle- and low-income countries. The rapid review will follow the PRISMA guidelines, and the literature search will be performed across five relevant databases, as well as various government websites to scan for grey literature. Articles will be screen against pre-agreed inclusion/ exclusion criteria, and data will be extracted using a pre-defined charting table.

**Ethics and Dissemination:** All data rely on secondary, publicly available data sources; therefore no ethical clearance is required. Upon completion, the results of this study will be disseminated via the Imperial College London Community and published in an open access, peer-reviewed journal.

**Article Summary:** *Strengths and Limitations of this Study:*

- This systematic review protocol is the first to focus on a longitudinal analysis of pandemic preparedness policy development across low, middle and high income country settings
- This protocol and subsequent review benefit from increased transparency, a systematised strategy (PRISMA), and a reduction in the risk of bias, through publication in an open access journal
- This review will also capture grey literature - studies published outside peer-reviewed journals
- This review protocol and methodology is not as robust as systematic reviews, therefore will lack some of the robustness often associated will classical systematic reviews

**Registration Number:** Open Science Framework: 10.17605/OSF.IO/VKA39

## Introduction

A pandemic is a disease that spreads across multiple countries, and can last for several months or years. Many pandemics are caused by infectious diseases, and often affect the elderly and immunocompromised (1); now, with the increasing rates of air travel, globalization and urbanization, such infectious diseases are able to spread widely and rapidly across immune-naive populations (2).

During any pandemic, governments and policy makers must continue to update their communities, and enforce certain policies, to ensure proper public health precautions are being followed (2). With new diseases emerging and the rapid development of connective technologies, pandemic response plans require flexibility and revision to ensure effectiveness (3). This dynamism allows pandemic preparedness and response to adapt, as more epidemiological, diagnostic and clinical information become available (2). Non-pharmaceutical interventions (NPI) are imperative at a time when pharmacological intervention is largely absent and/ or experimental. And while NPIs can be recommended, or indeed enforced, by local and national government, successful population uptake of policies such as mask-wearing, curfews and international travel restrictions is likely a function of previous sensitisation to infectious disease outbreaks and the democratic environment.

Pandemics strain national economies and healthcare systems (4). Unemployment rates may increase, as many businesses are forced to furlough or close, while the healthcare system may struggle with bed and ICU capacity, medication and human clinical resources (4).

High-income countries often have access to a greater number of financial and specialist human resources, which enables the effective and rapid scale-up of surveillance capacity (4). At the same time, economic provision, in the form of furlough or ‘helicopter money’, remains at the disposal of central banks, given the high demand for government-back bonds and a strong currency (5). Often, large parts of the workforce are employed in the services economy, many of whom are able to rely on technology to work remotely, and therefore commit to social distancing policies with greater ease and less negative financial and public health impact (6).

Low- and middle-income countries (LMICs) are broadly affected by the same basic epidemiological and population characteristics as high-income countries, however varying transmission dynamics, influenced by factors such as population density, under-funded surveillance and fewer health facilities, can exacerbate transmission (7). That said, LMICs have both greater experience in managing infectious disease outbreaks, and younger populations, which can alter the shape of epidemics.

### Rationale

The 20^th^ century was subjected to Spanish Influenza, Asian Influenza and Hong Kong Influenza, while the 21^th^ century saw the emergence of SARS, H1N1 and COVID-19. To varying degrees, these pandemics shaped the political, economic and social status quo of each affected nation. Importantly, each pandemic also altered pandemic preparedness and response, captured by peer-reviewed articles that evaluated government actions during the height of each event.

Accordingly, this study aims to review and analyse the evidence for effective pandemic preparedness policy, via a rapid systematic review of retrospective policy evaluations using the PRISMA guidelines (10). In so doing, this study aims to develop a roadmap of the significant policy changes that have occurred to-date, stratified by high, middle and low-income nations, in order to better inform future pandemic preparedness policy.

### Objectives

The objectives of this rapid systematic review are to:

1. identify published academic evaluations of national pandemic response across low-, middle- and high-income countries;
2. synthesise qualitative and quantitative data relating to the evaluation of national and international pandemic preparedness;
3. compare and contrast pandemic preparedness across contexts, and develop suitable recommendations.

## METHODS

### Study Design

This rapid systematic review will be conducted in accordance with the Preferred Reporting Items for Systematic Reviews and Meta-Analyses (PRISMA) guidelines. A rapid systematic review is best suited for this study, given time and funding constraints, and the depth of the subject matter.

### Search Strategy

Searches will be conducted by EL using the medical subject headings, free text terms and Boolean operators detailed in Appendix 1. The following databases will be searched to identify appropriate peer-reviewed literature: Embase, Web of Science, PubMed, PMC and Medline. Results will be screened and reference lists from included articles will also be searched.

### Study Selection Criteria

All studies published after 1918, which coincides with the date of the Spanish Flu, will be included.

Initial selection will be based on review of titles and abstracts alone. Duplicates will be removed, followed by a second stage of screening of the full text of each document.

Grey literature sources, such as government pandemic preparedness and response documents, will be obtained from government websites, specifically from the Ministry of Health Department. International pandemic response and preparedness documents will be found via the World Health Organisation (WHO) website.

### Eligibility Criteria

Countries will be defined as low, middle- and high-income by their Gross National Income (GNI), as defined by the World Bank. The World Bank uses the Atlas Method to calculate GNI, and adjusts for exchange rate (11). As of 2019, the GNI of a high-income country is defined as > $12,375, a middle-income country is defined as $1,026-$12,375 and a low-income country is defined as < $1,026 (12). As this rapid systematic review will analyse pandemics since 1918, each country will be identified as low, middle- and high-income, based on their GNI at the time.

Studies will be excluded if they do not include data from the six pandemics analysed. This review discusses government response and preparedness towards pandemics, particularly response using non-pharmaceutical interventions. Grey literature will also be searched and analysed. Both qualitative and quantitative studies will be included. The Hudden Matrix, adapted for pandemic influenza preparedness, will be used to justify successes and failures outlined by each included article (13). Other criteria inclusions and exclusions are detailed in Table 1.

**Table 1.**
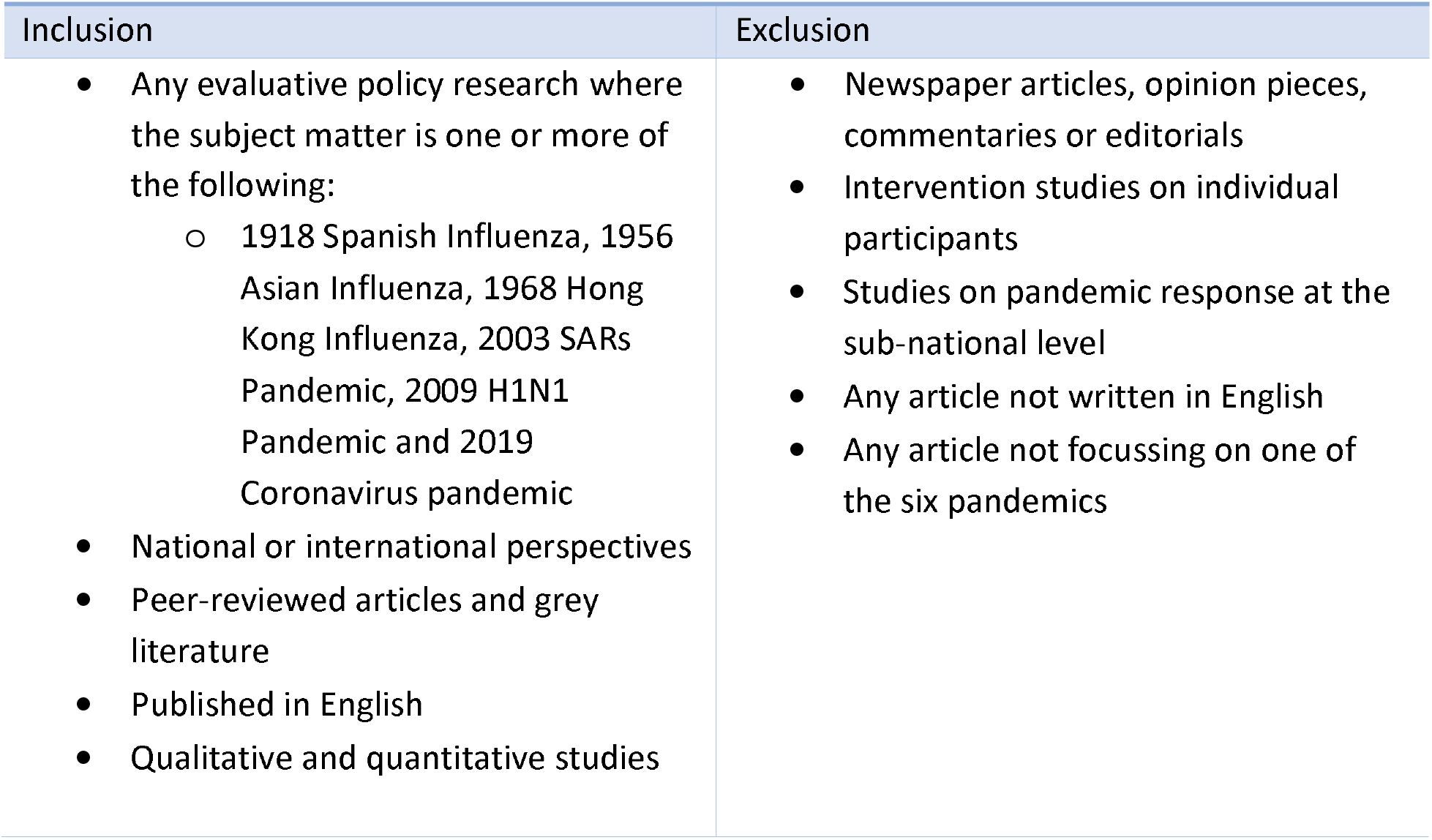
Inclusion and Exclusion Criteria.

### Study Selection

All the studies will be sourced via online databases, and a detailed record will be kept of keywords, search constructs and results. Duplications will be removed. Abstracts, and then the full text of each included study, will be screened against the inclusion and exclusion criteria. EL will screen each of the studies, with support from HT and LB. There is a risk of publication bias using the aforementioned strategy, which will be partially offset by searching grey literature and the reference list of each of the included articles.

### Data Extraction

Data extraction will be performed by EL using the charting template in Appendix 2, which has been adapted for this particular study, to ensure standardisation. HT and LB will review the data extraction process weekly.

### Outcomes

Both qualitative and quantitative outcomes will be included, although this rapid review will employ a thematic analysis.

### Patient and Public Involvement

Patients, nor the public, will be involved at any stage during this research.

## DISCUSSION

This rapid systematic review focuses on preparedness and response at the national and international level. It will analyse the evolution of preparedness using previous pandemic case studies to create a roadmap to inform future national and international pandemics preparedness and response plans.

## Data Availability

Availability of data and materials: This protocol has been registered at the following: Open Science Framework: DOI 10.17605/OSF.IO/VKA39. All data are available within the manuscript.

https://osf.io/vka39/

## Declarations

### Consent for publication

Not applicable.

### Availability of data and materials

This protocol has been registered at the following: Open Science Framework: DOI 10.17605/OSF.IO/VKA39. All data are available within the manuscript. Competing interests The authors declare that they have no competing interests.

## Author contributions

LB conceived this scoping review; LB, HT and EL collaborated in developing the aims and objectives, search strategy, and in selecting which methods to use to conduct this review.

## Acknowledgements

The authors would like to thank the School of Public Health at ICL for their continued support.

## Appendices

**Appendix 1.**
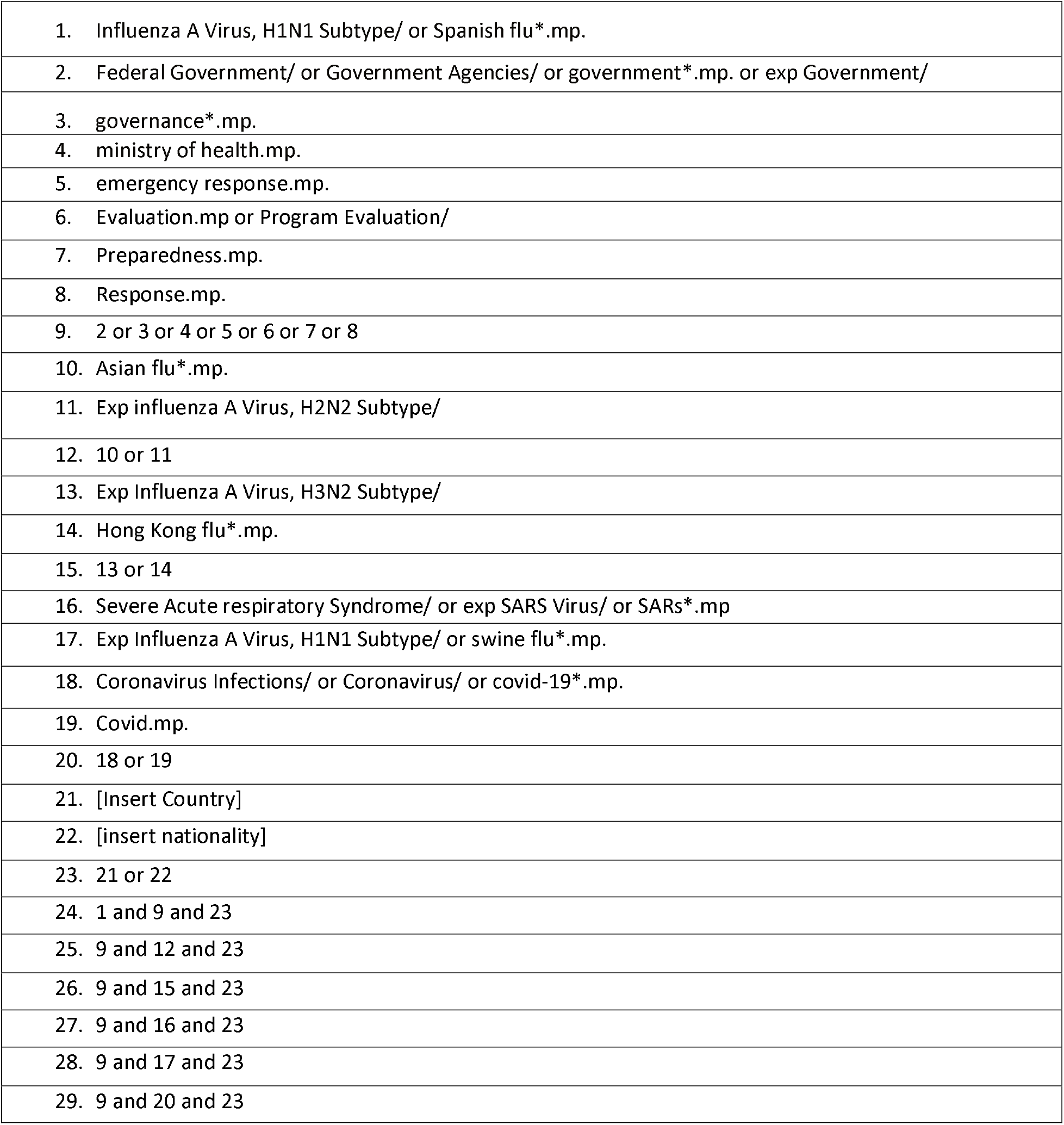
Search Strategy for Medline.

**Appendix 2.**
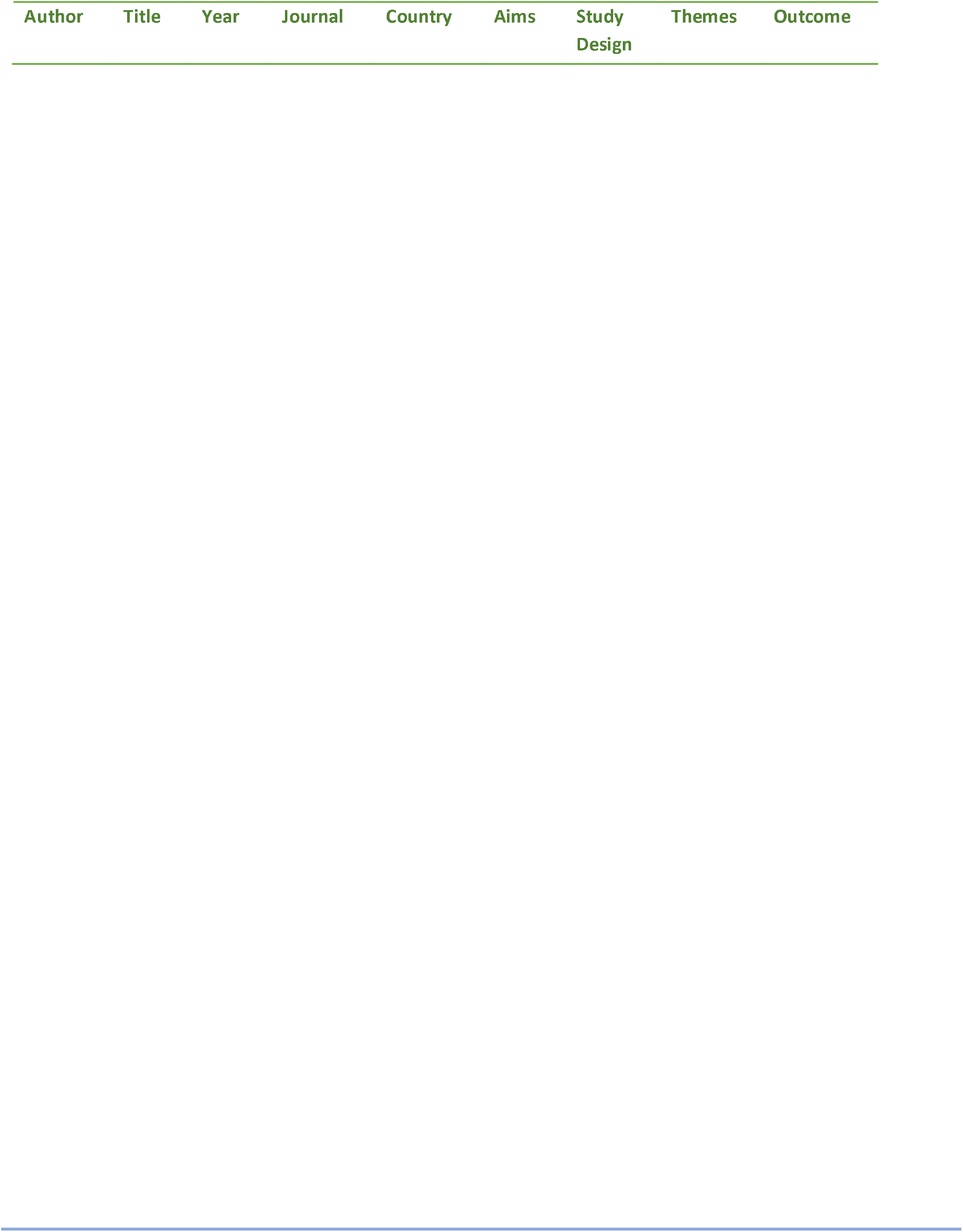
Charting Table.

